# Discovering Hidden Links: Harnessing Similarity Network Fusion to Reveal Common Clusters in Healthy Aging, Mild Cognitive Impairment, and Dementia

**DOI:** 10.1101/2025.04.02.25325137

**Authors:** Taeko Bourque, Peter Zhukovsky, Cassandra Morrison, John A. E. Anderson, the Alzheimer’s Disease Imaging Initiative (ADNI)

## Abstract

Cognitively Unimpaired (CU), Mild Cognitive Impairment (MCI), and Alzheimer’s Disease (AD) are clinical labels used to categorize degrees of cognitive impairment in the aging brain. Older adults experience brain changes associated with aging and cognitive decline at different ages and progress at varying rates, leading to heterogeneous patterns of cognitive decline. As a result, people within the same diagnostic category often exhibit significant differences in cognitive abilities and brain functions. The goal of the present study was to investigate how data-driven categories map onto diagnostic categories. Therefore, using data from 515 participants in the Alzheimer’s Disease Neuroimaging Initiative (ADNI) database (adni.loni.usc.edu) and a multivariate clustering approach, Similarity Network Fusion (SNF), we combined brain imaging features (cortical thickness average, surface area, and volume) with cognitive measures (Alzheimer’s Disease Assessment Scale-Cognitive, Mini-Mental State Exam, Montreal Cognitive Assessment, Clinical Dementia Rating) in older adults who had a clinical diagnosis of CU, MCI, or AD. We identified four data-driven groups that overlap diagnostic classifications and feature more similar neural and cognitive profiles. The current study provides evidence that more nuanced, data-driven approaches can reveal commonalities in the etiology and underlying neurobiology of individuals across traditional categories of CU, MCI, or AD. These results may lead to more focal therapies and a better understanding of who is at risk for converting to dementia, allowing for earlier detection and treatment of cognitive decline.

## Introduction

Dementia affects approximately 50 million people worldwide (World Health Organization, 2019), and this number is expected to double every 20 years (Prince et al., 2015). This fast-growing global health concern can be devastating not only to affected individuals but also to their families, caregivers, and economies. Dementia is used to label impaired functioning due to cognitive decline, with Alzheimer’s Disease (AD) accounting for approximately two-thirds of dementia diagnoses (Jalbert et al., 2008; Karantzoulis & Galvin, 2011). Despite the advent of new amyloid-clearing drugs, treating dementia remains very challenging. Therefore, identifying those at high risk for dementia in the earliest stages of the disease and targeting risk factors to slow down the progression of the disease is critical to improving clinical outcomes. Mild Cognitive Impairment (MCI), also known as prodromal (i.e., the beginning of a condition, when symptoms first appear) AD, is an early stage of AD and is used to label cognitive decline that is detectable by clinical measures but does not cause impaired functioning in daily tasks (Baldeiras et al., 2008; Dawe et al., 1992; Lee, 2023; Linn et al., 1995; Petersen et al., 1999). Individuals with MCI may remain stable over time or transition to AD; the conversion rate is approximately 10 to 18% (Davatzikos et al., 2011; Landau et al., 2010; Peterson et al., 2009). Determining who will transition from MCI to AD or dementia remains challenging, particularly because of the heterogeneity of disease progression in neurodegenerative disorders; brain functions decline at different ages and rates in different people (Dunn et al., 2022; Maheux et al., 2023; Young et al., 2018). For example, some individuals with differing clinical labels (i.e., CU, AD, MCI) may have more similar neural and cognitive profiles than indicated by current categorical classifications.

Recent studies highlight this heterogeneity and emphasize the need for new approaches to move the field forward (Bermejo-Pareja & Del Ser, 2024; Dunn et al., 2022; Maheux et al., 2023; Young et al., 2018). For example, Maheux et al. (2023) used disease progression models to predict the progressive decline from patients across all stages of AD. Disease progression models use computational and statistical methods to analyze patients’ various clinical and biomarker data and learn the variability of disease progression. Maheux et al. used this method on 4600 participants spanning four continents and found that AD Course Map provided a robust and generalizable predictive method. Although MCI has been described as a prodromal stage to AD, this topic is controversial because of the heterogeneity in presentation of both disorders and the unclear progression of MCI to AD (Bermejo-Pareja & Del Ser,2024; Dawe et al., 1992; Lee, 2023; Thompson & Hodges, 2002). The psychometric boundaries for an MCI diagnosis are evolving and not well standardized (Lee, 2023; Swallow, 2020; Vega & Newhouse, 2014). Indeed, revised MCI criteria have been criticized as blurring the contrast between MCI and mild AD, which some argue comes at the cost of a potential AD diagnosis (Lee, 2023; Morris, 2012). Practitioners may also be concerned with the clinical and social factors of a diagnosis (Swallow, 2020).

Data-driven clustering methods may reveal subgroups of older adults that exhibit greater similarities in brain or behavior that may be lost with traditional diagnostic categories. For example, promising work by Jacobs et al. (2021) used Similarity Network Fusion (SNF; Wang et al., 2014) to demonstrate the possibility of studying new data-driven groups of children that transcended traditional diagnostic classification of Autism, Obsessive-Compulsive Disorder (OCD) and Attention-Deficit/Hyperactivity Disorder (ADHD). Through SNF, participants are grouped based on their similarities to other participants within and across different data types, resulting in clusters of individuals who share the greatest resemblance. Grouping individuals based on shared neural and cognitive similarities may reveal more nuanced classifications of individuals on the spectrum of normal to abnormal aging, which are not captured by the current diagnostic classifications.

The objective of this study was to use SNF (Jacobs et al., 2021; Wang et al., 2014) to refine traditional diagnostic categories of cognitive impairment in the aging brain by identifying groups of aging individuals that are most similar to each other based on brain and cognitive scores. We hypothesized that SNF would reveal distinct groups of older adults with varying profiles of cognitive decline, where individuals within each cluster would share more similarities in brain imaging and behavioral traits, regardless of their clinical diagnostic status. Furthermore, we anticipated that certain individuals categorized as cognitively unimpaired (CU) might display preliminary signs of neurocognitive deterioration, potentially increasing their susceptibility to future decline. Our findings revealed four clusters of participants with more nuanced divisions across the spectrum of cognitive decline.

## Methods

Data used in preparing this article were obtained from the Alzheimer’s Disease Neuroimaging Initiative (ADNI) database (adni.loni.usc.edu). The ADNI was launched in 2003 as a public-private partnership led by Principal Investigator Michael W. Weiner, MD. The primary goal of ADNI has been to test whether serial magnetic resonance imaging (MRI), positron emission tomography (PET), other biological markers, and clinical and neuropsychological assessment can be combined to measure the progression of mild cognitive impairment (MCI) and early Alzheimer’s disease (AD). Participants were selected from ADNI-1, ADNI-GO, ADNI-2, and ADNI-3 cohorts. The study received ethical approval from the review boards of all participating institutions. Written informed consent was obtained from participants or their study partners.

### Participants

There were a total of 5411 participant data points across all phases (i.e., ADNI1, ADN1-GO, ADNI2, ADNI3, ADNI4) and cohorts (i.e., CU, AD, MCI) in the ADNI database. However, as the ADNI database contains longitudinal data (i.e., multiple data for the same participant across time), we only included the most recent and complete entry for each participant. Additionally, participants were excluded from our analyses if they had missing MRI imaging data or missing cognitive data for either the ADAS-Cog, MMSE, MoCA, or CDR. These criteria resulted in the data from 515 participants (all from either ADNI 1 or ADNI 3) being used for our study.

### Measures

#### Demographics

Participants reported their sex (F, M), age (years), and level of education (number of years of formal education completed).

##### Depression

Depression was measured as the total raw score of the Geriatric Depression Scale (GDS; Yesavage & Sheikh, 1986). The GDS is a self-report measure designed to assess depressive symptoms in older adults. There are 15 yes/no questions, with yes questions scoring one point. A score of five points indicates possible depression, with higher scores indicating greater depressive symptom severity.

##### Vascular Cognitive Impairment

The Hachinski Ischemic Scale (HIS; Hachinski et al., 1975) is used to differentiate between types of dementia, such as Alzheimer’s disease or vascular dementia. The HIS has eight questions with a possible score of zero, one, or two points depending on the severity of symptoms. Scores of four or less points are indicative of Alzheimer’s disease, whereas scores of seven or more points are indicative of vascular dementia.

#### Clinical/Cognitive Assessments

Four cognitive measures were selected to represent clinical features of neurodegeneration: the Alzheimer’s Disease Assessment Scale-Cognitive (ADAS-Cog; Rosen et al., 1984), the Mini-Mental State Exam (MMSE; Folstein et al., 1975), the Montreal Cognitive Assessment (MoCA; Nasreddine et al., 2005), and the Clinical Dementia Rating (CDR; Morris, 1993). Total raw scores for each cognitive measure were used in the SNF analysis. These measures are all used to assess cognitive impairment, including questions that involve orientation to the current time and place, ability to recall and recognize words from lists, naming objects, performing multi-step commands, problem-solving, repeating items forwards and backwards, copying shapes, or general conversation ability. The MoCA is particularly effective at early detection of MCI, whereas the MMSE is easier and more suited to assessing the severity and progression of cognitive impairment when screening for Dementia. The ADAS-Cog is used to assess the level of cognitive dysfunction in AD and the CDR is commonly used to assess the severity of cognitive dysfunction in AD or other dementias.

#### P-tau 181

The tau protein phosphorylated at threonine 181 (P-tau181) is biomarker in cerebrospinal fluid where increased levels are specific for AD. P-tau 181 for the ADNI database was analyzed by the Single Molecule array technique, using an in-house assay developed in the Clinical Neurochemistry Laboratory, University of Gothenburg, Sweden (Karikari et al., 2020).

#### Image Analysis

Cortical volume, thickness, and surface area were derived from T1-weighted images using the Desikan-Killiany Atlas in FreeSurfer (6.0). These derivatives were taken from the ADNI database. Imaging metrics for cortical thickness average, surface area (divided by intracranial volume), and cortical volume (divided by intracranial volume) were used as brain-based input features in the SNF analysis.

### Procedure

#### MRI Acquisition Parameters

MRI data were acquired using 3T scanners, providing higher resolution images, typically around 1.0 mm x 1.0 mm x 1.0 mm, with a similar or slightly larger FOV depending on the scanner and specific protocol used. The 3T imaging protocols also included T1-weighted MP-RAGE sequences, which offer improved signal-to-noise ratio and contrast, beneficial for detailed structural analysis. Further details on the MRI acquisition parameters and protocols can be found at ADNI MRI Methods (https://adni.loni.usc.edu/methods/mri-tool/mri-analysis/).

#### Similarity Network Fusion (SNF) and Spectral Clustering

Similarity Network Fusion (Wang et al., 2014) is a computational method for data integration that leverages common and complementary information in different types of data. Figure 1 illustrates how this method iteratively processes individual similarity networks (for data type), thus pruning or strengthening connections between participants for a final, fused participant similarity network representing all data types. After calculating a robust similarity network matrix, we used spectral clustering to determine the data clusters (i.e., groups).

**Figure 1.**
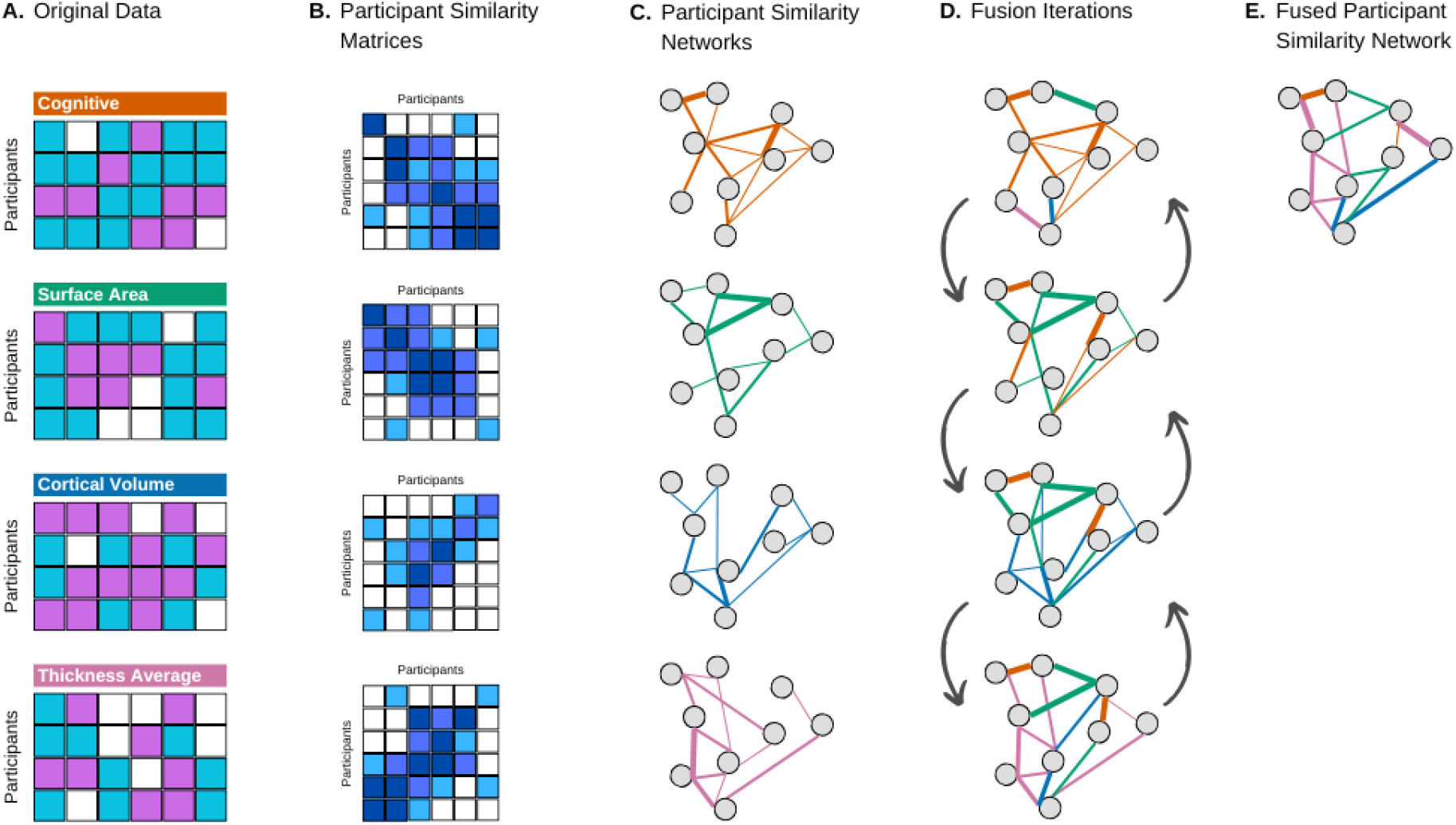
Overview of the Similarity Network Fusion Approach (Wang et al. 2014)

### Analyses

All analyses were performed using R Statistical Software (v4.3.1; R Core Team 2023) and closely followed code published by Jacobs et al. (2021). The scripts used for analyses have been made available at https://github.com/TaekoBourque/SNF-ADNI. We combined various brain imaging features (i.e., cortical thickness average, surface area, and cortical volume) with cognitive measures (i.e., Alzheimer’s Disease Assessment Scale-Cognitive - ADAS-Cog; Mini-Mental State Exam - MMSE; Montreal Cognitive Assessment - MoCA; Clinical Dementia Rating - CDR) in older adults with a clinical diagnosis of CU, MCI, or AD. These structural imaging and cognitive data types were integrated using the *SNFtool* package (v2.3.1; Wang et al., 2021).

## Results

### Data Integration and Cluster Determination

First, Euclidean distance matrices are calculated for each data type (i.e., cognitive, cortical thickness average, surface area, cortical volume). Following Wang et al. (2014) and Jacobs et al. (2021) methods, we chose the following hyperparameters: the number of neighbors was set to 18, the normalization hyperparameter to 0.8, and the number of iterations to 10. Subsequently, Euclidean distance matrices, where lower values indicate greater similarity, are converted into affinity matrices (i.e., similarity matrices) which capture the pairwise similarities between data points. These similarity matrices form the basis of participant similarity networks, where each node represents an individual, and edges encode similarity strength based on each data type. Finally, a fused participant similarity network is created by iteratively updating each network with information from other networks. At each iteration, individual networks are updated by exchanging structural information with the others, ensuring that both shared and modality-specific similarities are retained. To assess the contribution of each feature to the fused similarity structure, we compute Normalized Mutual Information (NMI), which quantifies the degree of overlap between a similarity matrix based on one feature and the final fused matrix (based on all features). NMI ranges from 0 (no mutual information) to 1 (perfect correlation), with higher values indicating a greater influence of that feature on the overall participant similarity structure. Table 1 shows the top 35 SNF input features ranked by Normalized Mutual Information (NMI; see supplemental materials for all 210 SNF input features).

**Table 1.** The Top 35 Ranking Model Features (by NMI) Contributing to Participant Similarity and Corresponding Statistical Summaries of Data-Driven and Diagnostic groups.

| Rank | Top Contributing Features | Normalized Mutual Information | Data-Driven Groups |  |  | Diagnostic Groups |  |  |
| --- | --- | --- | --- | --- | --- | --- | --- | --- |
|  |  |  | F value | pFDR | Eta squared | F value | pFDR | Eta squared |
| 1 | right lingual SA | 0.340 | 0.024 | 0.893 | 0.006 | 1.240 | 0.338 | 0.005 |
| 2 | right lateral orbitofrontal SA | 0.289 | 1.410 | 0.290 | 0.005 | 1.520 | 0.270 | 0.006 |
| 3 | right medial orbitofrontal SA | 0.279 | 12.200 | 0.001 | 0.033 | 6.680 | 0.004 | 0.027 |
| 4 | right middle temporal SA | 0.238 | 5.250 | 0.039 | 0.029 | 7.170 | 0.002 | 0.029 |
| 5 | left rostral anterior cingulate TA | 0.103 | 3.790 | 0.081 | 0.009 | 2.260 | 0.147 | 0.009 |
| 6 | left bankssts TA | 0.097 | 32.000 | 0.000 | 0.080 | 16.500 | 0.000 | 0.064 |
| 7 | right bankssts TA | 0.091 | 41.700 | 0.000 | 0.092 | 20.500 | 0.000 | 0.078 |
| 8 | left caudal middle frontal TA | 0.080 | 26.400 | 0.000 | 0.069 | 16.800 | 0.000 | 0.065 |
| 9 | left lateral occipital TA | 0.076 | 7.210 | 0.015 | 0.017 | 1.670 | 0.237 | 0.007 |
| 10 | right supramarginal TA | 0.073 | 36.200 | 0.000 | 0.090 | 15.800 | 0.000 | 0.061 |
| 11 | right rostral anterior cingulate TA | 0.071 | 2.800 | 0.129 | 0.019 | 3.180 | 0.068 | 0.013 |
| 12 | right pericalcarine TA | 0.068 | 0.466 | 0.553 | 0.010 | 2.070 | 0.169 | 0.009 |
| 13 | right rostral middle frontal TA | 0.067 | 9.280 | 0.006 | 0.026 | 4.950 | 0.016 | 0.020 |
| 14 | right caudal middle frontal TA | 0.067 | 22.700 | 0.000 | 0.065 | 15.100 | 0.000 | 0.059 |
| 15 | right lateral occipital TA | 0.065 | 15.300 | 0.000 | 0.037 | 5.950 | 0.007 | 0.024 |
| 16 | right cuneus TA | 0.063 | 2.140 | 0.184 | 0.010 | 1.840 | 0.204 | 0.007 |
| 17 | left entorhinal TA | 0.061 | 135.000 | 0.000 | 0.320 | 87.100 | 0.000 | 0.260 |
| 18 | left superior frontal TA | 0.061 | 21.600 | 0.000 | 0.064 | 13.500 | 0.000 | 0.053 |
| 19 | left rostral middle frontal TA | 0.058 | 23.900 | 0.000 | 0.055 | 12.300 | 0.000 | 0.048 |
| 20 | left cuneus TA | 0.055 | 0.000 | 0.996 | 0.005 | 1.040 | 0.400 | 0.004 |
| 21 | left inferior parietal TA | 0.053 | 31.400 | 0.000 | 0.082 | 17.500 | 0.000 | 0.067 |
| 22 | left precentral TA | 0.053 | 13.600 | 0.001 | 0.042 | 6.860 | 0.003 | 0.028 |
| 23 | MMSE | 0.052 | 307.000 | 0.000 | 0.640 | 212.000 | 0.000 | 0.470 |
| 24 | left pericalcarine TA | 0.052 | 0.592 | 0.499 | 0.011 | 1.010 | 0.407 | 0.004 |
| 25 | right pars triangularis TA | 0.051 | 7.300 | 0.015 | 0.020 | 4.910 | 0.016 | 0.020 |
| 26 | left superior temporal TA | 0.051 | 74.900 | 0.000 | 0.170 | 45.300 | 0.000 | 0.160 |
| 27 | right temporal pole TA | 0.049 | 49.800 | 0.000 | 0.140 | 28.800 | 0.000 | 0.110 |
| 28 | right superior temporal TA | 0.048 | 59.300 | 0.000 | 0.150 | 32.800 | 0.000 | 0.120 |
| 29 | left pars triangularis TA | 0.048 | 10.300 | 0.004 | 0.022 | 4.460 | 0.024 | 0.018 |
| 30 | right superior frontal TA | 0.047 | 18.100 | 0.000 | 0.049 | 6.490 | 0.004 | 0.026 |
| 31 | left insula TA | 0.047 | 33.400 | 0.000 | 0.110 | 19.500 | 0.000 | 0.074 |
| 32 | left lingual TA | 0.047 | 7.290 | 0.015 | 0.019 | 3.090 | 0.072 | 0.013 |
| 33 | right entorhinal TA | 0.047 | 101.000 | 0.000 | 0.270 | 68.500 | 0.000 | 0.220 |
| 34 | right frontal pole TA | 0.046 | 0.904 | 0.406 | 0.015 | 0.716 | 0.523 | 0.003 |
| 35 | left isthmus cingulate TA | 0.044 | 21.600 | 0.000 | 0.060 | 10.900 | 0.000 | 0.043 |

*Note.* SA (surface area); CV (cortical volume); TA (thickness average); NMI (Normalized Mutual Information); *pFDR* (False Discovery Rate adjusted p-value); MMSE (Mini-Mental State Exam).

To determine the optimal number of clusters, potential solutions ranging from two to ten clusters were evaluated using the fused similarity matrix. The analysis was repeated across combinations of nearest neighbors (*K* = 10–30) and affinity scaling parameters (*α* = 0.3–0.8). A four-cluster solution was the most consistently supported across the parameter combinations and was therefore selected for the final spectral clustering analysis. To assess the stability of the clustering solution, robust spectral clustering was performed using 1,000 resampling iterations, with 80% of participants randomly sampled during each iteration. A silhouette plot (see Supplementary Materials) was used to examine the similarity between participants within a given group compared to participants in all other groups. Silhouette widths varied; Group 1 (0.87) and Group 3 (0.88) had strong cluster structures, Group 2 (0.52) had a good cluster structure, and Group 4 (0.45) had a slightly weaker structure. Overall, the average silhouette width was 0.55, indicating good cluster structure. Network Visualization (see Figure 2, Panel A) was used to illustrate the participant similarity networks in a way that highlights participants’ data-driven group assignment and their clinical diagnosis.

**Figure 2.**
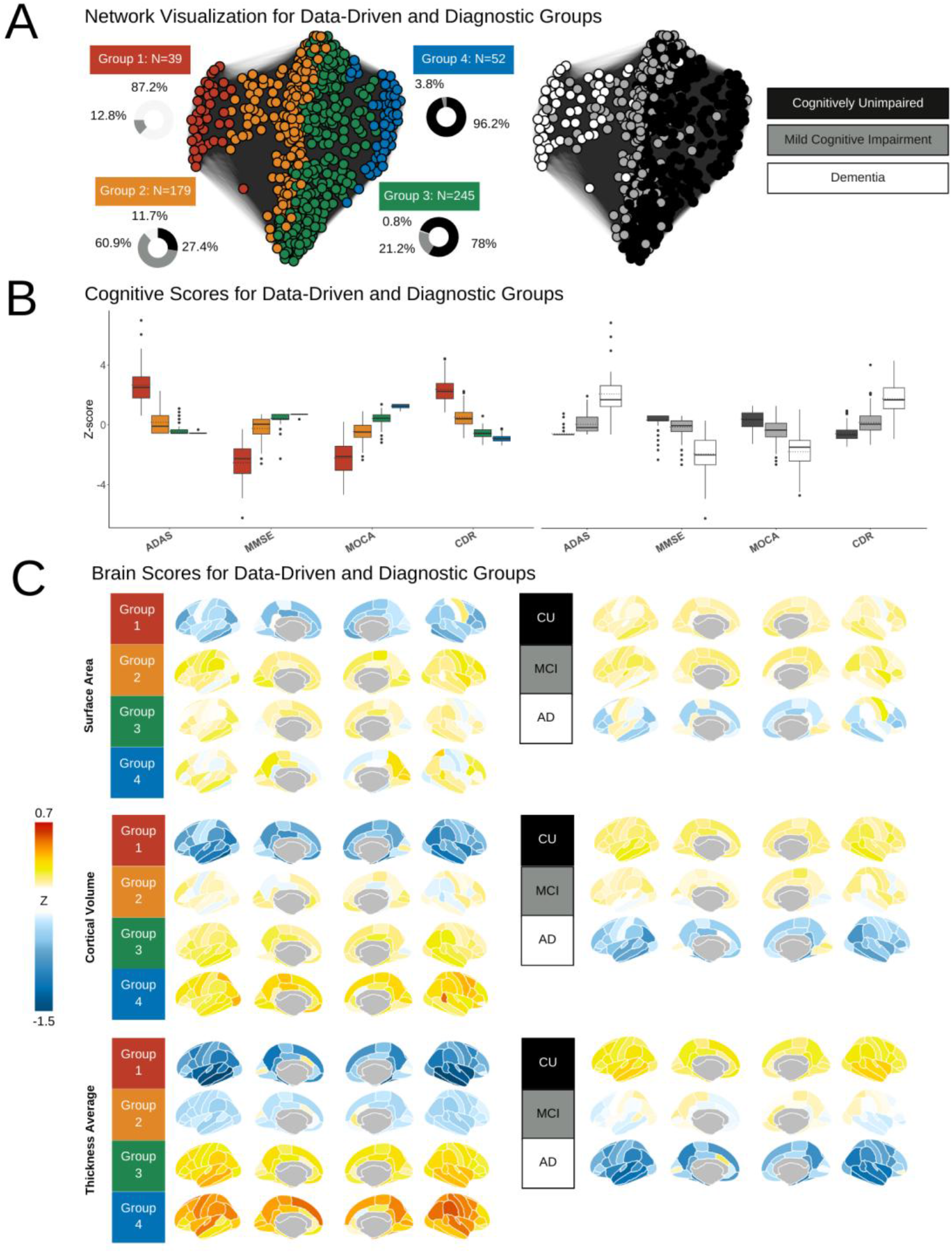
Relative Participant Similarities, Cognitive Scores, and Brain Scores for Data-Driven and Diagnostic Groups. *Note*. A) Data-driven groups (left) and those same groups colored by clinical diagnoses (right). B) Boxplots representing the cognitive z-scores for data-driven (left) and diagnostic (right) groups. ADAS = Alzheimer’s Disease Assessment Scale-Cognitive; MMSE = Mini-Mental State Exam; MoCA = Montreal Cognitive Assessment; CDR = Clinical Dementia Rating. C)

### SNF Shows Four Groups Spanning a Gradient of Cognitive and Neural Severity

Based on participant similarity matrices determined by 210 model features, 4 distinct groups were identified (see Figure 2). The four data-driven groups featured an average silhouette width of 0.55, indicating a good cluster structure. These four clusters spanned a gradient from most severely affected cognitively and neurologically (group 1) to relatively unaffected (group 4). The top 10 model features with the highest NMI scores included the surface area of the right lingual, right lateral orbitofrontal, right medial orbitofrontal, right middle temporal, as well as the cortical thickness average of the left rostral anterior cingulate, left and right bankssts, left caudal middle frontal, left lateral occipital, and right supramarginal gyrus. As per Jacobs et al. (2021), we chose to feature the top 35 features contributing to participant similarity (see Table 1). Of these top features, the MMSE was the only model feature other than additional cortical thickness average and surface area regions.

#### Comparisons Among Data-Driven and Diagnostic Groups

Following Jacobs et al. (2021) we conducted separate one-way ANCOVAs among data-driven groups and among diagnostic groups (see Table 1) to provide a standardized effect size estimate (eta-squared) of group distinctions for all 210 model features that contributed to participant similarity. The p-value was corrected for multiple comparisons using a false discovery rate (pFDR) of 5%.

Mirroring the approach to plotting cognitive scores, z-scores for surface area, cortical volume, and thickness average are plotted in brain space using *ggseg* (DK atlas). CU = Cognitively Unimpaired; MCI = Mild Cognitive Impairment; AD = Alzheimer’s Disease.

*Note*. SA (surface area); CV (cortical volume); TA (thickness average); NMI (Normalized Mutual Information); *pFDR* (False Discovery Rate adjusted p-value); MMSE (Mini-Mental State Exam).

#### Comparison of Top Contributing Features Between Data-Driven and Diagnostic Groups

Separate one-way ANCOVAS showed a significant main effect of group (after accounting for the effects of age, gender, and depression) across the top 35 ranking model features. These top features included 30 measures of cortical thickness average, 4 measures of surface area, 1 cognitive measure, and 0 measures of cortical volume (see Figure 3 and Table 1). Effect sizes were typically smaller, and fewer were significant in diagnostic groups relative to data-driven groups.

**Figure 3.**
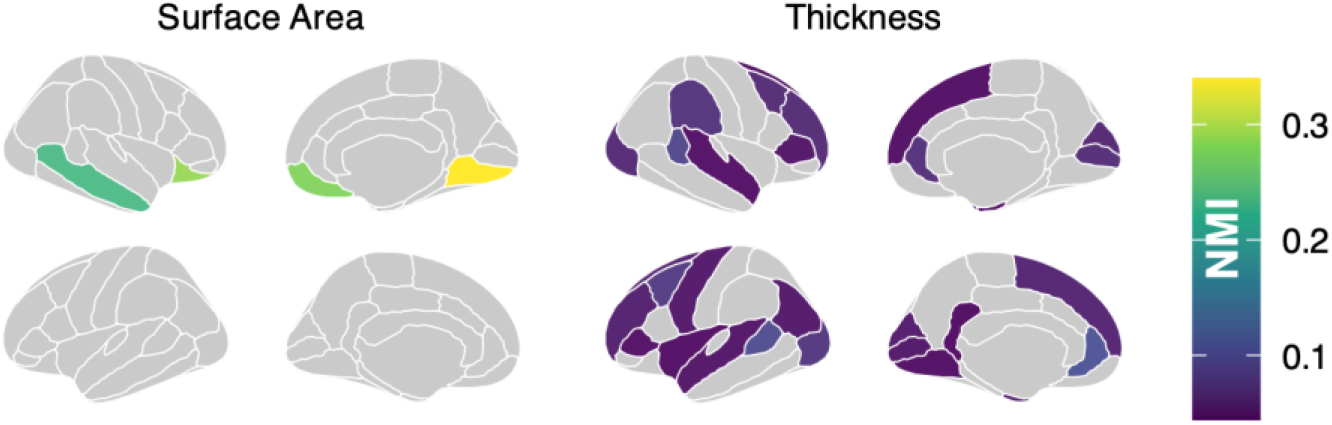
Representation of Top Ranking SNF Features in Brain-Space

#### Demographic Differences

Sex, age, education, depression, and Hachinski score were not included as input features in the SNF analysis. They were instead used to examine if there were demographic features that differed between the groups. Table 2 shows the descriptive statistics of the data-driven groups (see Supplemental Materials for descriptive statistics of diagnostic groups).

**Table 2.** Descriptive Statistics of Data-Driven Groups.

|  | **Group 1**<br>**(n = 39)** | **Group 2**<br>**(n = 179)** | **Group 3**<br>**(n = 245)** | **Group 4**<br>**(n = 52)** | **$\chi^2$, df = 3** | ***p*** |
| --- | --- | --- | --- | --- | --- | --- |
|  | M (SD) | M (SD) | M (SD) | M (SD) |  |  |
| Sex (F) | 16 (41%) | 53 (30%) | 134 (55%) | 37 (71%) | 41.161 | <0.001 |
| Age (years) | 72.84 (7.39) | 73.10 (7.14) | 70.53 (6.20) | 68.18 (5.68) | 30.391 | <0.001 |
| Education (years) | 15.53 (2.82) | 16.05 (2.68) | 16.76 (2.39) | 17.33 (2.26) | 18.15 | <0.001 |
| Depression Score | 1.92 (1.63) | 1.99 (2.45) | 1.27 (1.77) | 0.81 (0.97) | 23.12 | <0.001 |
| Hachinski Score | 0.67 (0.84) | 0.61 (0.75) | 0.49 (0.58) | 0.31 (0.61) | 10.763 | 0.013 |

*Note.* Descriptive statistics for sex are represented as the number of females (with the percentage of the group they represent in brackets), and a Chi-squared test was used to test for group differences. For age, education, depression, and Hachinski ischemic scale score, descriptive statistics are represented as means (with standard deviations in brackets), and Kruskal-Wallis tests were used to test group differences.

#### Biomarker Differences

Age and p-tau differences between groups are shown in Figure 4. Panel A displays age differences across data-driven and diagnostic groups. Groups 3 and 4, which exhibit fewer clinical symptoms, tend to be younger compared to groups that are more severely affected by neurodegeneration and cognitive impairment. Panel B illustrates p-tau 181 levels by group, showing that Group 1 has higher concentrations of p-tau than Group 3, while Group 2 also has elevated p-tau levels relative to Group 3. No significant differences in p-tau levels were observed between Groups 3 and 4, suggesting that the early decline detected by the SNF model may precede an increase in p-tau concentrations.

**Figure 4.**
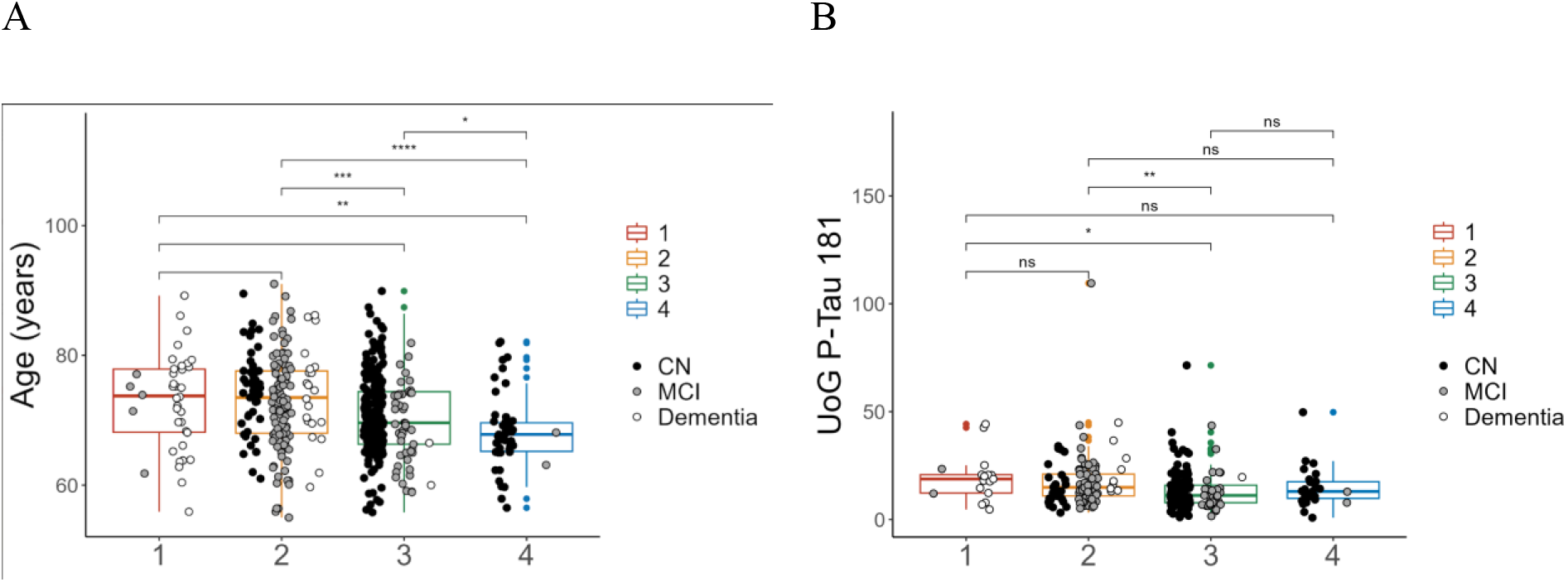
Age and P-Tau Differences Between Groups. *Note*. ns: p > 0.05, *: p <= 0.05, **: p <= 0.01, *: p <= 0.001, **: p <= 0.0001. All *p*-values are FDR corrected for multiple comparisons.

### Can SNF Reveal At-Risk Groups of Cognitively Unimpaired Older Adults?

Our clustering solution shows a significant overlap with diagnostic labels for dementia and MCI. Specifically, 87% of individuals in Group 1 were diagnosed with dementia, while the rest were diagnosed with MCI. In Group 2, 60.9% were diagnosed with MCI, 11.7% with dementia, and 24.7% were cognitively unimpaired. In the cognitively unimpaired group, we see the most notable differences between diagnostic categories and model results. This group was divided into two: Group 4 (which contained 96.2% CU), which showed no signs of impairment on neuropsychological tests or brain indices, and Group 3 (which contained 78% CU, and 21.2% MCI diagnoses), which exhibited early, statistically significant declines in these metrics compared to Group 4. A possible explanation is that participants in Group 3 might be showing signs of incipient neuropathology and cognitive decline. As the present research is cross-sectional, further research is required to determine whether these individuals would indeed transition from CU to MCI.

## Discussion

Neurodegenerative diseases like AD and MCI can have highly heterogeneous symptoms and progression. Traditional diagnostic categories (i.e., CU, MCI, AD) oversimplify this complexity and pull focus to changes of kind rather than of degree. This study employs Similarity Network Fusion (SNF) and spectral clustering to integrate brain imaging and cognitive measures and address the challenge of reconciling diagnostic categories with a person’s underlying aetiology and performance on neuropsychological tests. Analyses revealed novel, data-driven groups of participants from the ADNI data, grouping individuals who shared more similarities with each other than their diagnoses would suggest. We predicted that transdiagnostic groups of individuals would emerge based on brain and neuropsychological features, better reflecting their current cognitive status than their diagnoses alone. We also anticipated that some individuals classified as cognitively unimpaired (CU) would be identified as having indicators of incipient neuropathology. Early identification of individuals who present normally on neuropsychology screening instruments, but whose neurological profiles suggest they are declining may allow for more rapid intervention and personalized treatments.

In line with our prediction that our data fusion approach would reveal different groupings than the diagnostic labels, our similarity network fusion analysis identified ***four*** distinct groups of individuals, with each group showing highly similar brain and neuropsychological profiles, varying in degree of neurodegeneration and cognitive function. These findings refine traditional diagnostic categories by revealing more nuanced divisions across the spectrum of cognitive decline. Importantly, the groups spanned a gradient of severity, generally aligning with traditional categories (i.e., CU, MCI, AD), but with surface area, cortical thickness, and MMSE emerging as the most important distinguishing features. Group 1 included the most severely impaired individuals, and the majority had AD diagnoses. Group 2 was mostly participants with MCI, with some AD participants. Group 3 was mostly CU, with some MCI; this group had early signs of neuropathology. And finally, Group 4 was largely CU, and individuals with minimal cognitive impairment.

Interestingly, our analysis favored a four-group solution over the traditional three-category grouping. This additional group split the CU and MCI groups at the extreme end of the x-coordinates producing Group 3 (see Figure 2). This finding suggests that many individuals categorized as ‘cognitively unimpaired’ may carry early signs of MCI, nearly imperceptible by routine cognitive screening. Future studies should investigate whether subjective cognitive decline measures, such as the Cognitive Change Index, might further differentiate these groups. Increasing evidence supports the idea that individuals with subjective cognitive decline, previously labeled ‘the worried well,’ may actually be in the early stages of dementia (Jessen et al., 2014; Mendonça et al., 2016; Rabin et al., 2017), and thus, Group 3 may be the group that is most likely to progress to cognitive decline. Our approach emphasizes a continuum model of cognitive and neuropathological decline rather than a sudden shift that aligns with diagnostic thresholds.

Cortical thickness, especially in medial and lateral temporal regions and in precuneus were the most important features for distinguishing groups and was the best predictor of cognitive decline along a gradient consistent with the known progression of AD (Schwarz et al., 2016). Surface area and volume metrics were useful for differentiating the most severely affected individuals from those unaffected but was less useful for discerning early changes. Of the cognitive measures, only the MMSE appeared among the top SNF features highlighting its diagnostic utility, despite its limited sensitivity to early decline. Future studies may wish to see if other measures such as the MoCA provide additional insights into differentiating CU from early progressors. Our findings also support prior research suggesting that cortical thinning is a more reliable marker of disease progression than surface area changes (Bakkour et al., 2013; Dickerson et al., 2009; Williams et al., 2023).

Our approach highlights the importance of fusion techniques for offering insights which transcend single modalities and offer a sensitive approach to re-thinking traditional diagnostic categories. Specifically, we suggest that traditional diagnostic categories may miss key early variations in disease progression, particularly when traditional neuropsychological screening tools are used as gates through which a patient must pass before other, complementary diagnostic features are included. Traditional diagnostic categories, especially those that rely on standard neuropsychological screening tools as a primary diagnostic step, may overlook crucial early signs of disease progression and fail to capture the full spectrum of disease variability. High levels of heterogeneity among participants within diagnostic categories also pose a problem for clinical trials and could cause them to fail to detect subtle but important effects or require vastly more participants to boost power (Reed et al., 2021; Shapiro & Usner, 2012). Using SNF-derived groups as a starting point in clinical trials could thus help to improve clinical trial sensitivity and provide more homogeneous subgroups for targeted interventions. Others have used a similar approach to fuse DNA, and molecular measures with cognitive decline in the RUSH dataset (Yang et al 2023) and revealed two subtypes associated with diffuse amyloid plaques. Using these approaches may thus offer more comprehensive views of the aging brain.

Our findings mark an important step toward personalized medicine by demonstrating how advanced MRI features, integrated with machine learning, can provide clinicians with tools to detect early cognitive decline. Cognitive and neural decline exist on a continuum, and detecting early signs of deterioration remains a challenge, particularly in patients who do not fall neatly into a single diagnostic category. Surface area and cortical thickness—integral to our model—are essential for capturing these early changes, but without MRI, they are difficult to assess.

### Limitations and Future Directions

Previously, work by Jacobs et al. (2021), emphasized a lack of overlap between clinical diagnosis and biological and neuropsychological burden (Jack Jr. et al., 2018; Sperling et al., 2011) - this was not the case for the present results. Our results revealed a similar pattern between data-driven and clinical groups; however, our model emphasized a gradient and appears more sensitive to early change. This is a relatively homogeneous sample - it would be good to validate the findings in another large open-access dataset (e.g., RUSH, ONDRI). Future work could extend input features to include genetic and protein factors (Yang et al., 2023), explore how these profiles are affected by cognitive reserve or risk factors (Stern 2002, 2012), or investigate whether individuals transition from one group to another. Emerging technologies such as ultra-low-field MRI (0.055T) along with novel tools like SynthSR offer promising, cost-effective solutions for monitoring brain health longitudinally, even in clinical settings. These advances, coupled with rapid cortical thickness estimation algorithms like FASTsurfer, could allow real-time integration of patient data into predictive models like the one outlined in our study, aiding in early diagnosis and the tracking of cognitive decline progression (Shen et al., 2021).

## Conclusion

The current study adds to recent work highlighting the heterogeneity within diagnostic groups (Bermejo-Pareja & Del Ser,2024; Dunn et al., 2022; Maheux et al., 2023; Young et al., 2018. We identified new data-driven groups that overlap diagnostic classifications and feature more similar profiles of brain and behavior. Cognitive impairment is a continuum, and the sharp delineations of diagnostic categories (i.e., CU, MCI, AD) do not allow for a more nuanced interpretation. The current study provides evidence that more nuanced, data-driven approaches can allow for better representation of a participant’s brain and behavior profile that could allow for earlier detection and treatment.

## Data Availability

Data used in the preparation of this article were obtained from the Alzheimer's Disease Neuroimaging Initiative (ADNI) database (http://www.loni.ucla.edu/ADNI). The scripts used for analyses have been made available at https://github.com/TaekoBourque/SNF-ADNI.

https://github.com/TaekoBourque/SNF-ADNI

## Acknowledgments

Data collection and sharing for the Alzheimer’s Disease Neuroimaging Initiative (ADNI) is funded by the National Institute on Aging (National Institutes of Health Grant U19 AG024904). The grantee organization is the Northern California Institute for Research and Education. In the past, ADNI has also received funding from the National Institute of Biomedical Imaging and Bioengineering, the Canadian Institutes of Health Research, and private sector contributions through the Foundation for the National Institutes of Health (FNIH) including generous contributions from the following: AbbVie, Alzheimer’s Association; Alzheimer’s Drug Discovery Foundation; Araclon Biotech; BioClinica, Inc.; Biogen; Bristol-Myers Squibb Company; CereSpir, Inc.; Cogstate; Eisai Inc.; Elan Pharmaceuticals, Inc.; Eli Lilly and Company; EuroImmun; F. Hoffmann-La Roche Ltd and its affiliated company Genentech, Inc.; Fujirebio; GE Healthcare; IXICO Ltd.; Janssen Alzheimer Immunotherapy Research & Development, LLC.; Johnson & Johnson Pharmaceutical Research & Development LLC.; Lumosity; Lundbeck; Merck & Co., Inc.; Meso Scale Diagnostics, LLC.; NeuroRx Research; Neurotrack Technologies; Novartis Pharmaceuticals Corporation; Pfizer Inc.; Piramal Imaging; Servier; Takeda Pharmaceutical Company; and Transition Therapeutics. Metabolomics Consortium (ADMC) will include the following language: Data collection and sharing for this project was funded by the Alzheimer’s Disease Metabolomics Consortium (National Institute on Aging R01AG046171, RF1AG051550 and 3U01AG024904-09S4).

## Author Note

We have no known conflict of interest to disclose. This project was supported by an NSERC Discovery Grant (DGECR-2022-00309) and a Canada Research Chair (Tier II, CRC-2020-00174) to JAEA.

